# When to be vaccinated? What to consider? Modelling decision-making and time preference for COVID-19 vaccine through a conjoint experiment approach

**DOI:** 10.1101/2021.06.05.21258416

**Authors:** Samson W.H. Yuen, Ricci P.H. Yue, Bobo H.B. Lau, Cecilia L. W. Chan, Siu-Man Ng

**Affiliations:** Department of Government and International Politics, Hong Kong Baptist University, Hong Kong; Department of Geography, University of Hong Kong; Department of Counselling and Psychology, Hong Kong Shue Yan University, Hong Kong; Department of Social Work and Social Administration, University of Hong Kong

## Abstract

How do citizens choose COVID-19 vaccines, and when do they wish to be vaccinated? A choice-based conjoint experiment was fielded in Hong Kong to examine factors that shape citizens’ preference toward COVID-19 vaccines and their time preference to be vaccinated, which is overlooked in extant literature. Results suggest people are most concerned about vaccines’ efficacy and severe side-effects, and that cash incentives are not useful in enhancing vaccine appeal. The majority of respondents show low intention for immediate vaccination, and many of them want to delay their vaccination. Further analysis shows that their time preference is shaped more by respondent characteristics than vaccine attributes. In particular, confidence in the vaccine, trust in government, and working in high-risk professions are associated with earlier timing for vaccine uptake. Meanwhile, forced COVID testing would delay vaccination. The findings offer a novel view in understanding how people decide whether and when to receive new vaccines, which have pivotal implications for a head start of any mass vaccination programs.

**Highlights:**

- People are most concerned about vaccines’ efficacy and severe side-effects when choosing COVID-19 vaccines
- Cash incentives are not useful in enhancing vaccines’ appeal
- Time preference of vaccination is shaped more by respondent characteristics than vaccine attributes
- Forced COVID testing might delay vaccination decision

## Introduction

The COVID-19 pandemic has caused over 130 million infections and 3 million deaths around the world while heavily devastating the global economy. ^1^ Although non-pharmaceutical interventions, such as draconian social distancing policies, have substantially contained the spread of the pandemic, there has been a consensus among governments that a mass vaccination program is needed for citizens to develop immune responses against infections, which can in turn alleviate the burden for the public health system and pave the way toward herd immunity.

Several COVID-19 vaccines have been given authorizations for emergency use by COVAX and national governments. Corresponding mass vaccination programs are gradually rolled out in many developed countries. Researchers, meanwhile, have also begun to examine vaccine acceptance or vaccine hesitancy, along with the factors that significantly impact vaccine intention. In June 2020, a cross-sectional survey in 19 countries reported a 71.5% acceptance of COVID-19 vaccine in the most affected countries (Lazarus *et al*., 2020). A more recent 32-country survey conducted from October to December 2020 showed that vaccine acceptance in key countries ranged from 38% to 98%, displaying a geographical disparity in vaccine intention (Wouters *et al*., 2021). Previous efforts have also identified government trust (Lazarus *et al*., 2020) risk perception (Sherman *et al*., 2020), and demographic background (Malik *et al*., 2020; Paul *et al*., 2021) as determinants of vaccination intention.

However, understanding the degree of vaccine uptake intention, and what contributes to such intention, are not sufficient for formulating an effective vaccine policy. Health policy-makers also need to know how vaccine characteristics and citizen-level characteristics influence vaccination choice and, mostly importantly, time preference. While most existing studies focus on COVID-19 vaccine intention (Detoc *et al*., 2020; Harapan *et al*., 2020; Rhodes *et al*., 2020), limited research has been conducted on how vaccine characteristics shape the choice of vaccine and the preferred timing for vaccination. The latter is particularly important because intention is merely a general inclination. It does not tell us how people choose between vaccines and the timing at which they want to be vaccinated. For example, intention to vaccinate is likely to be highly contingent on the efficacy of a given vaccine. Also, although some people indicate that they have no intention in being vaccinated, it may be that they are simply delaying their plans to get vaccinated because they want to wait out and see if the vaccines are safe.

In February 2021, Hong Kong approved its first COVID-19 vaccine based on the recommendation of an advisory panel formed by medical experts. The government eventually decided to procure three vaccines – Sinovac, Fosun Pharma/BioNTech and Oxford AstraZeneca – which vary in manufacturing origins, efficacy and potential side-effects according to clinical trials. While officials hoped that a successful mass vaccination scheme would control the pandemic at a time when the city was facing a fourth wave infection, the public appeared to be hesitant toward the vaccines. An earlier local study based on a survey conducted in summer 2020 projected an overall vaccine acceptance rate of 37.2% (Wong *et al*., 2021). A subsequent study even indicated that only 13.1% of the citywide population had the intention to take up vaccination at its earliest availability. The discrepancy also hinted that Hong Kong citizens have different time preferences toward vaccination (Yu *et al*., 2021). Questions over whether the residents intend to take COVID-19 vaccine, which vaccines are preferred, and how to implement the vaccine programme were hotly debated before the start of the vaccination program.

To address the gap, we conducted a large-N online survey in Hong Kong in January 2021 prior to the start of the vaccination program. Preferences toward different vaccine attributes and time preference for vaccination are measured through a conjoint model embedded in the survey. Furthermore, their time preferences on receiving vaccines with different values are estimated against government trust, risk perceptions and demographic backgrounds of the respondents. This study can help inform mass vaccine program planning and the allocation of vaccines with different characteristics for different countries.

## Materials and Methods

We conducted a large-N cross-sectional survey in Hong Kong on adults aged above 18 between 22 January 2021 and 28 January 2021. The survey is the latest wave of a four-wave panel survey that aims to understand citizens’ psychological conditions under the COVID-19 pandemic. A total of 2,733 respondents were recruited within the survey period using a non-probability sampling frame. A significant proportion of respondents were enlisted from the previous wave of the panel survey, while new respondents were also added to the panel through snowballing from existing respondents and the networks of NGOs. Given the emergent nature of the inquiry, non-probability sampling is the quickest way in which we can collect a large-enough sample for conducting survey experiments. Moreover, while data collection has been restricted by the severity of the pandemic and social distancing policy, we were able to circumvent the restrictions by conducting the survey through Qualtrics, an online survey platform. Participants who completed the survey were reimbursed a HKD50 worth of supermarket coupon for their time. According to Orme (2006), the equation for estimating the sample size of conjoint analysis is:

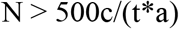

Where N is the minimum sample size; c is the largest number of attribute levels; it is the number of attributes tested; and a is the number of choices given in each task. This suggested a minimum survey population of 208 people. Our sample size clearly exceeded this requirement.

## Conjoint model

Conjoint analysis is a survey-based technique that helps to determine how people value different attributes that constitute a particular product, service or policy. It has been widely adopted in marketing research for measuring consumer preferences (Wittink & Cattin, 1989; Raghavarao *et al*., 2010); but has recently been extended to public policy and political science (Hainmueller *et al*., 2014). Although the technique has many variants, a common feature is that they ask respondents to choose from and/or rate hypothetical profiles that combine multiple attributes. This design allows researchers to estimate the relative causal influence of each attribute level on the resulting choice, which makes it a unique method to unravel the multidimensional considerations before choice-making.

Our conjoint experiment simulates the decision-making process of Hong Kong citizens in choosing between COVID-19 vaccines. Respondents were presented with two vaccine profiles, each characterized by six pre-selected attributes: 1) vaccine cost/subsidy, 2) vaccine efficacy, 3) protection scheme to compensate for severe side-effects, 4) likelihood of mild side-effects (e.g. swelling, fever, chills and/or tiredness), 5) likelihood of severe side-effects (e.g. severe allergic reaction or other life-threatening reactions), and 6) queuing time after registering for the injection. These attributes were chosen to approximate the information available to citizens who are considering whether to vaccinate and what vaccines to select, if they are given a choice. Each attribute can take on multiple levels. For instance, there are four levels in vaccine efficacy, which are randomized to characterize each profile across pairings. Table 1 contains the list of attribute levels. There are in total 6,000 unique vaccine profiles produced by the conjoint, which give us a wide range to assess which attributes are potentially influential.

**Table 1.**
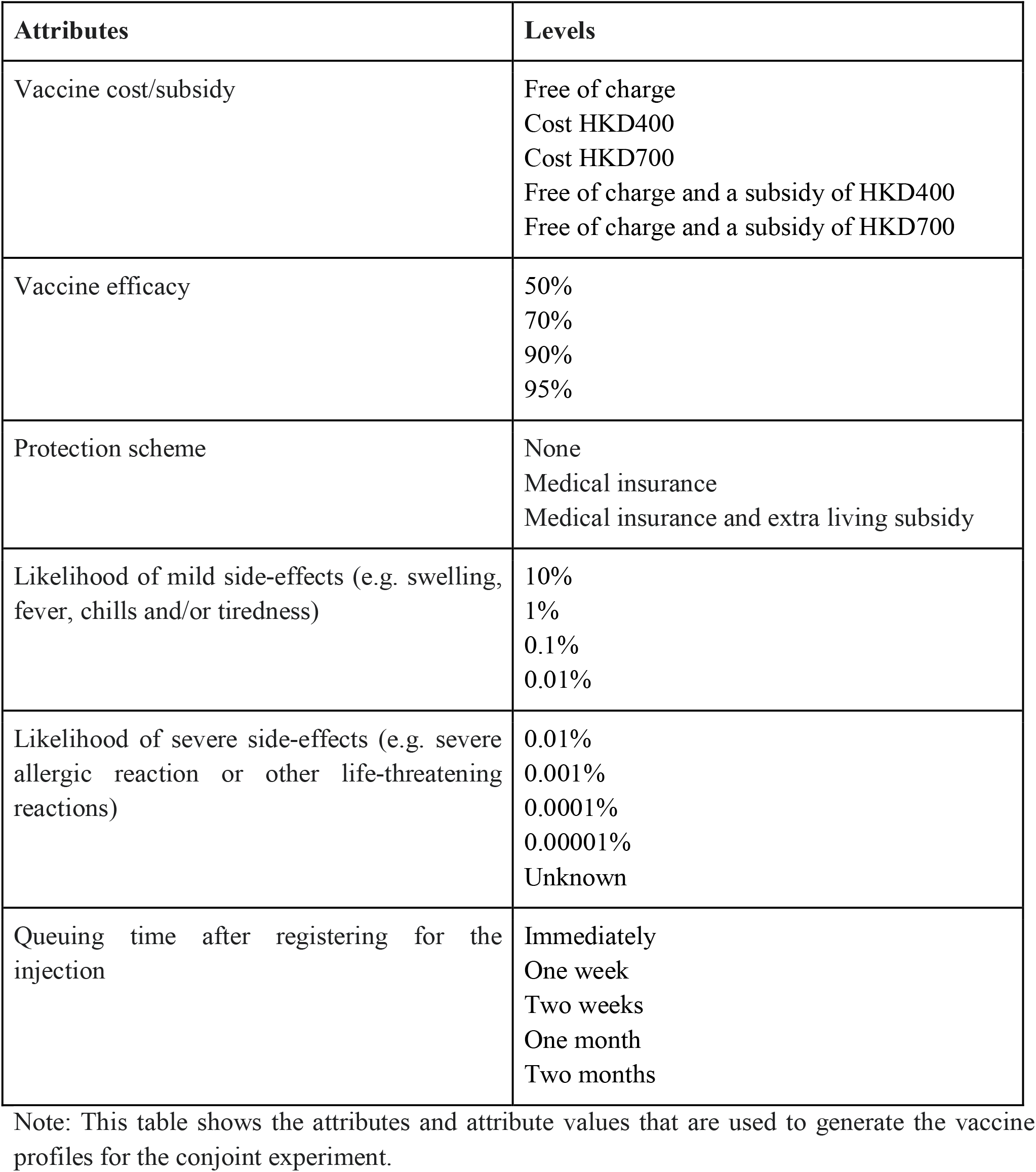
Attributes for Vaccine Profiles in Conjoint Experiment

Respondents were asked to provide a ‘forced choice’ from one of the two vaccine profiles, even though one might not have any intention to be vaccinated. To ensure enough statistical power, respondents were presented with three choice tasks. Order of the attributes is randomized for each respondent (but fixed across the three choice tasks) to minimize primacy and recency effect while easing the cognitive burden for respondents (Kumar & Gaeth, 1991; Chrzan, 1994).

For each choice task, besides indicating their choice of the preferred vaccine, respondents are further asked to state their time preference to be vaccinated for the choice that they made. Respondents can choose from ten options, each of which refer to the population quantiles in which they want to receive the injection. They range from the first 10% to be vaccinated among the population (1) to the last 10% to be vaccinated (10). This question is similar to the rating question that is often seen in conjoint experiments, in which respondents have to rate their degree of preference for a chosen profile. By reframing it as time preference, we are able to model how vaccine attributes, along with respondent-level attributes, influence the preferred vaccination timing.

## Results

### Modelling vaccine preference: Average marginal component effect

The statistical analysis is divided into two sections. In the first section we follow the approach proposed by Hainmueller *et al*. (2014) to estimate the average marginal component effects (AMCEs) of different attribute levels using their regression-based estimator. This can be achieved by running a linear regression of the choice outcome as the dependent variable on the sets of dummy variables for the attribute values. AMCEs can be interpreted as the effect sizes, which convey the expected change in the likelihood of selecting a particular profile when a given attribute level is compared with a chosen baseline attribute level.

The AMCE plot (Figure 1) yields the following results. Vaccine efficacy produces the largest spread of effect sizes. Vaccines with a 50% efficacy and those with a 70% efficacy are 30.7% and 16.9% less likely to be chosen than the baseline vaccine with a 95% efficacy. With respect to the probability of developing severe side-effects, a probability of 0.01%, 0.001% and 0.001% will make respondents 15.8%, 9.4% and 3.7% less likely to choose a vaccine compared with the baseline case of a 0.00001% probability. An unknown probability, meanwhile, will make respondents 22.6% less likely to choose that vaccine than the baseline. As for mild side-effects, a linear relationship similar to that of severe side-effects holds. The lower the likelihood of developing mild side-effects, the more preferable a vaccine will become.

**Figure 1.**
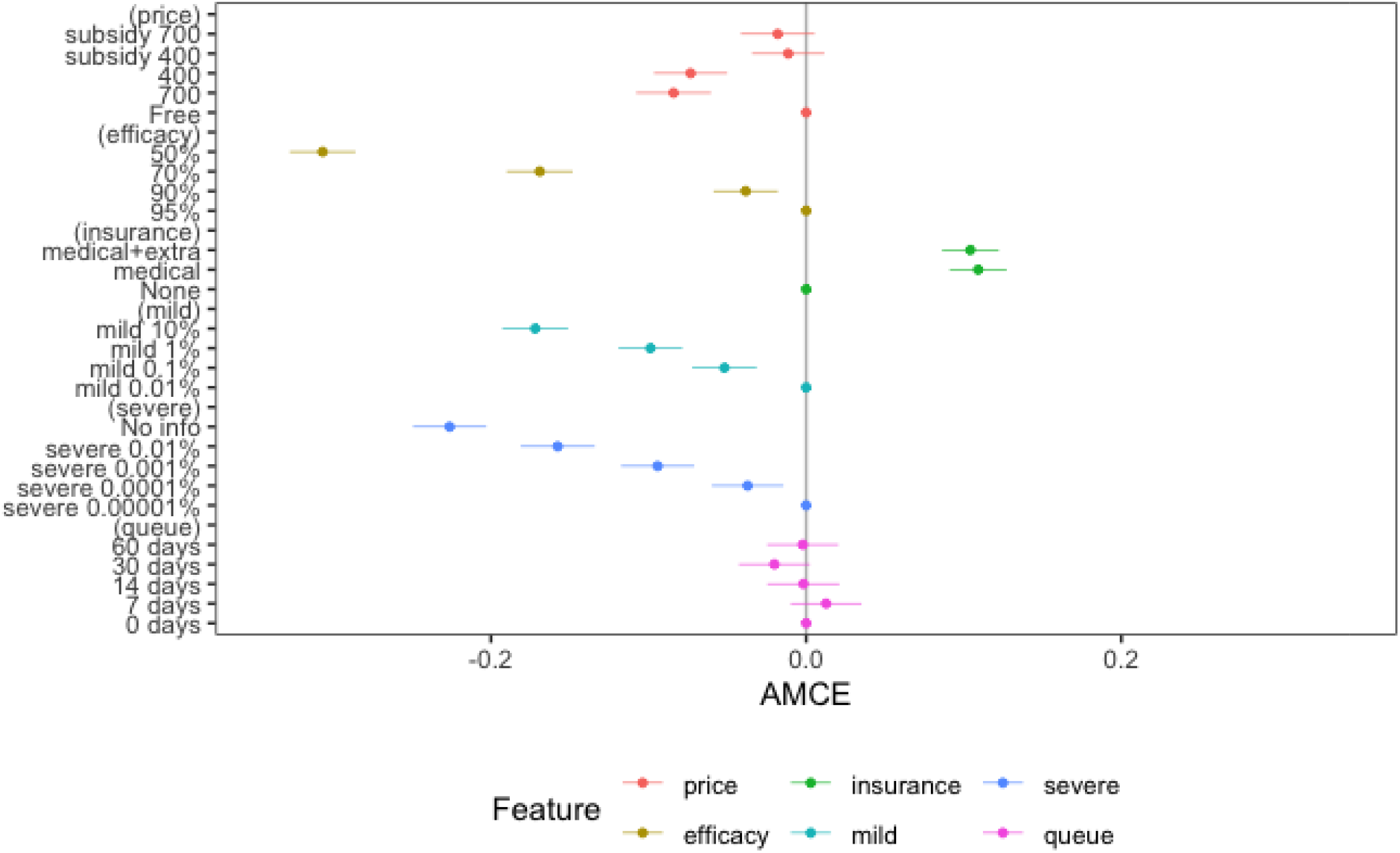
AMCE plot. Note: The dots indicate AMCE estimates, while the lines show the 95% confidence intervals for the AMCEs. The dots without confidence intervals denote the baseline categories. For instance, the first line from the top shows that vaccines that come with a subsidy of HKD700 are on average 8% less likely to be chosen than a vaccine that is free of charge, which is used as a baseline category. The coefficients are displayed in Table S2.

As for pricing, the results indicate that making people pay for vaccines will reduce their likelihood of choosing them. A vaccine that costs HKD700 and HKD400 respectively are 1.1% and 1.8% less likely to be chosen than a free vaccine. Interestingly, subsidizing vaccines also reduces the desirability of vaccines. A vaccine that comes with a subsidy of HKD400 and HKD700 are 8.4% and 7.3% less likely to be chosen than a free vaccine. This means that the more subsidy the government provides, the less desirable a vaccine becomes. One explanation is that subsidies may generate a negative signaling effect that guides people to think about the negative consequences of receiving the vaccine. Overall, this suggests that a free vaccine is most preferred by respondents.

On the other hand, providing medical insurance against severe side-effects will make a vaccine 10.9% more likely to be chosen than providing no such insurance. Interestingly, providing an extra living subsidy *does not* make vaccines more desirable than simply by providing medical insurance. Yet, it also will not make it markedly less desirable. Finally, there is no consistent and significant effect regarding queuing time after registering for vaccination. A wait of 7 and 14 days respectively is neither increasing nor decreasing the desirability of a vaccine as compared with no waiting time. However, a wait of 30 days will make a vaccine 2.0% less likely to be chosen than the baseline. Interestingly, if the wait prolongs to 60 days, the negative effect becomes negligible again.

### Time preference analysis

The second section focuses on vaccination time preference. While the conjoint experiment allows us to evaluate the relative importance of different vaccine attributes when making vaccination decisions, it does not tell us about the time at which respondents prefer to be vaccinated. The inclusion of the time preference question in each choice task provides an opportunity to examine this dimension. Our assumption is that people with different demographic backgrounds, government trust level and risk perception against COVID-19 would result in different vaccination time preference, based on earlier work on the determinants of vaccine uptake intention (Lazarus *et al*., 2020; Malik *et al*., 2020; Sherman *et al*., 2020; Paul *et al*., 2021). The summary descriptive statistics for these respondent-level characteristics are provided in Table S1.

We examine how vaccine attributes and respondent characteristics determine when they prefer to be vaccinated for the choice that they have made. Given that the time preference is a type of count data overdispersed with 10 (last 10% to be vaccinated) as the dominant time preference choice, we can model the data with zero-inflated negative binomial regression, which combines a negative binomial distribution and a logit distribution. Here, time preference, which is first recorded in reverse order, is specified as the dependent variable of the model. We include both the vaccine attributes in the conjoint experiment and a number of respondent-level characteristics as the independent variables in the negative binomial model. Meanwhile, we select two variables – 1) intention to vaccinate and 2) confidence in the vaccine – in the logit part of the model to predict the excess zeros. These variables are chosen because it is likely that respondents who do not want to be vaccinated and have low confidence in the vaccine will delay their vaccine preference as much as possible. We build the models hierarchically by adding blocks of variables on top of the others.

Figure 2 shows a histogram that shows the distribution of the time preferences for vaccination, which ranges from 1 to 10 (1 = first 10% to be vaccinated, 10 = last 10% to be vaccinated). Here time preference is also subsetted by respondents’ general intention to join the vaccination program. Majority (40.6%) of the choices are 10. The remaining responses are rather uniformly distributed along 1 to 9, but with a cluster toward the middle.

**Figure 2.**
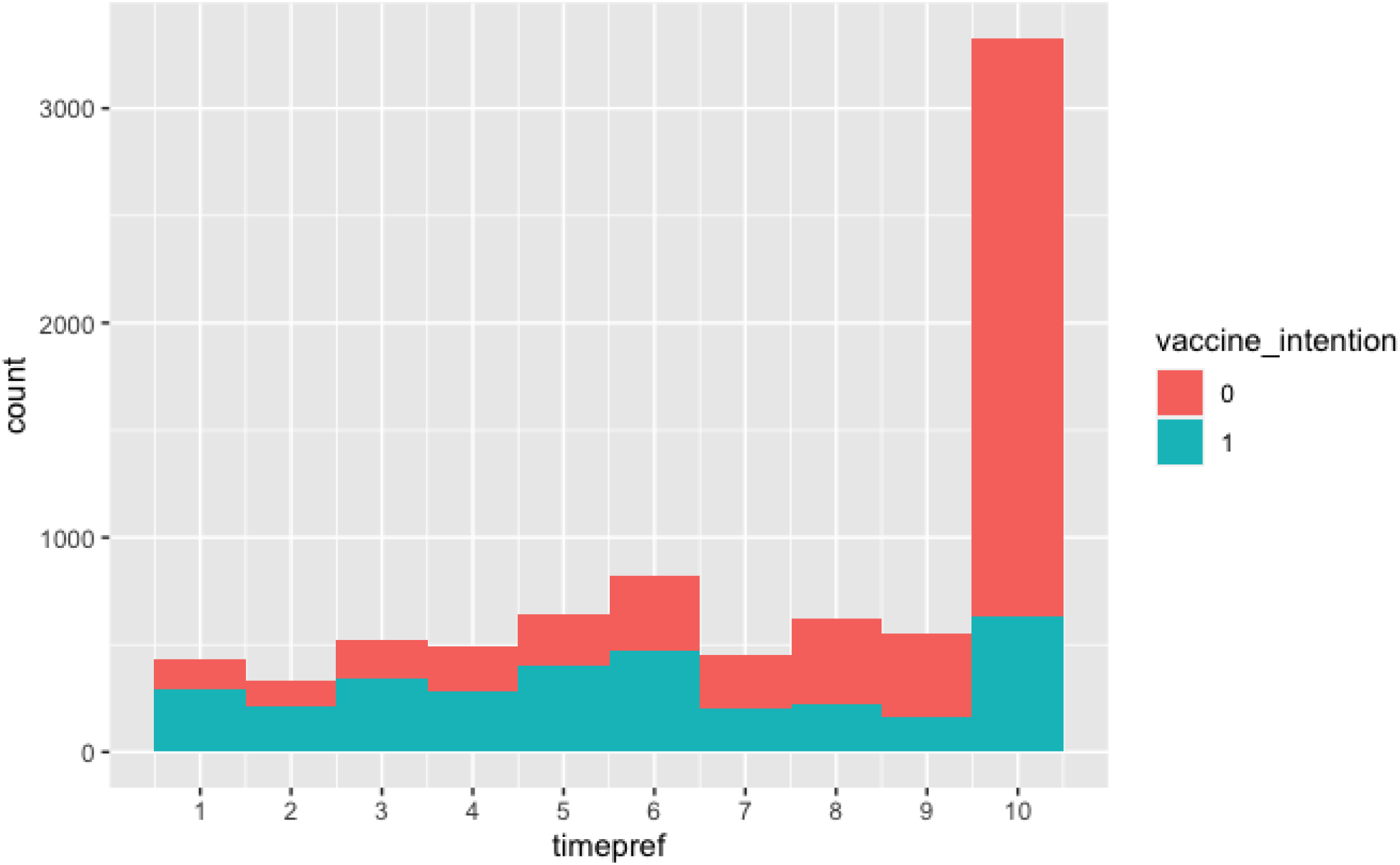
Time preference for vaccination.

The regression models are presented in Table 2. To interpret the results, one has to apply the natural exponential function to the estimates. For instance, the estimate for being male is 0.164 in the full model and the exponential function of that is 1.178. This means the average preferred vaccination timing for male respondents is 17.8% later than that for female respondents in terms of the population quantile.

**Table 2.** Zero-inflated negative binomial regression on time preference

| Results (negative binomial coefficients) from zero-inflated negative binomial regression |  |  |  |  |  |  |  |  |
| --- | --- | --- | --- | --- | --- | --- | --- | --- |
|  | Model |  |  |  |  |  |  |  |
|  | (1) | (2) | (3) | (4) | (5) | (6) | (7) | (8) |
| Age | 0.001**<br>(0.001) | 0.001**<br>(0.001) | 0.002***<br>(0.001) | 0.002***<br>(0.001) | 0.003***<br>(0.001) | 0.002**<br>(0.001) | 0.001**<br>(0.001) | 0.002**<br>(0.001) |
| Education | -0.057***<br>(0.006) | -0.056***<br>(0.006) | -0.057***<br>(0.007) | -0.055***<br>(0.007) | -0.051***<br>(0.007) | -0.047***<br>(0.007) | -0.046***<br>(0.007) | -0.047***<br>(0.007) |
| Sex | 0.167***<br>(0.016) | 0.167***<br>(0.016) | 0.166***<br>(0.016) | 0.168***<br>(0.016) | 0.171***<br>(0.016) | 0.162***<br>(0.016) | 0.161***<br>(0.016) | 0.159***<br>(0.016) |
| Income | 0.007**<br>(0.003) | 0.007**<br>(0.003) | 0.006*<br>(0.003) | 0.006*<br>(0.003) | 0.007**<br>(0.003) | 0.008**<br>(0.003) | 0.007**<br>(0.003) | 0.008**<br>(0.003) |
| Intention to be vaccinated | 0.212***<br>(0.017) | 0.214***<br>(0.017) | 0.214***<br>(0.017) | 0.217***<br>(0.017) | 0.222***<br>(0.017) | 0.149***<br>(0.020) | 0.154***<br>(0.020) | 0.084***<br>(0.025) |
| High risk jobs |  |  | 0.068***<br>(0.017) | 0.066***<br>(0.017) | 0.059***<br>(0.017) | 0.060***<br>(0.017) | 0.064***<br>(0.017) | 0.072***<br>(0.017) |
| Chronic illness |  |  | -0.017<br>(0.018) | -0.017<br>(0.018) | -0.023<br>(0.018) | -0.023<br>(0.018) | -0.020<br>(0.018) | -0.020<br>(0.018) |
| Reside with vulnerable persons |  |  | -0.003<br>(0.016) | -0.004<br>(0.016) | -0.001<br>(0.016) | -0.003<br>(0.016) | -0.005<br>(0.016) | -0.004<br>(0.015) |
| Perceived seriousness of COVID-19 |  |  |  | -0.0002<br>(0.005) | -0.002<br>(0.005) | 0.0003<br>(0.005) | 0.001<br>(0.005) | 0.002<br>(0.005) |
| Perceived risk of COVID infection |  |  |  | 0.006<br>(0.005) | 0.003<br>(0.005) | 0.002<br>(0.005) | 0.002<br>(0.005) | 0.002<br>(0.005) |
| Confidence in COVID prevention |  |  |  | -0.008*<br>(0.004) | -0.006<br>(0.004) | -0.012***<br>(0.005) | -0.012***<br>(0.005) | -0.012***<br>(0.005) |
| Financial impact |  |  |  |  | -0.004<br>(0.003) | -0.005<br>(0.003) | -0.005*<br>(0.003) | -0.005<br>(0.003) |
| Mental stress level |  |  |  |  | 0.015***<br>(0.003) | 0.016***<br>(0.003) | 0.016***<br>(0.003) | 0.016***<br>(0.003) |
| Trust in strangers |  |  |  |  |  | 0.008*<br>(0.004) | 0.006<br>(0.004) | 0.007<br>(0.004) |
| Trust in friends |  |  |  |  |  | -0.016***<br>(0.005) | -0.015***<br>(0.005) | -0.015***<br>(0.005) |
| Trust in medical workers |  |  |  |  |  | 0.005<br>(0.005) | 0.005<br>(0.005) | -0.005<br>(0.005) |
| Trust in government |  |  |  |  |  | 0.008*<br>(0.004) | 0.009**<br>(0.004) | 0.008*<br>(0.004) |
| Confidence in vaccine |  |  |  |  |  | 0.031***<br>(0.008) | 0.031***<br>(0.008) | 0.030***<br>(0.008) |
| Trust in vaccination program |  |  |  |  |  | 0.012*<br>(0.007) | 0.012*<br>(0.007) | 0.010<br>(0.007) |
| Tested |  |  |  |  |  |  | -0.034**<br>(0.020) | -0.256***<br>(0.050) |
| Confirmed case nearby (yes) |  |  |  |  |  |  | -0.085***<br>(0.027) | -0.002<br>(0.045) |
| Confirmed case nearby (Not sure) |  |  |  |  |  |  | -0.008<br>(0.037) | 0.051<br>(0.055) |
| Price: 700 |  | -0.031<br>(0.025) | -0.032<br>(0.025) | -0.032<br>(0.025) | -0.031<br>(0.025) | -0.033<br>(0.024) | -0.034<br>(0.024) | -0.031<br>(0.024) |
| Price: 400 |  | -0.027<br>(0.024) | -0.028<br>(0.024) | -0.028<br>(0.024) | -0.029<br>(0.024) | -0.033<br>(0.024) | -0.035<br>(0.024) | -0.036<br>(0.024) |
| Pricesubsidy_400 |  | -0.016<br>(0.023) | -0.017<br>(0.023) | -0.017<br>(0.023) | -0.016<br>(0.023) | -0.018<br>(0.023) | -0.017<br>(0.023) | -0.016<br>(0.023) |
| Pricesubsidy_700 |  | -0.043*<br>(0.024) | -0.043*<br>(0.024) | -0.043*<br>(0.024) | -0.040*<br>(0.024) | -0.044*<br>(0.024) | -0.044*<br>(0.024) | -0.040*<br>(0.024) |
| Efficacy: 90& |  | 0.010<br>(0.020) | 0.009<br>(0.020) | 0.010<br>(0.020) | 0.008<br>(0.020) | 0.008<br>(0.020) | 0.006<br>(0.020) | 0.006<br>(0.019) |
| Efficacy: 70% |  | 0.005<br>(0.021) | 0.002<br>(0.021) | 0.001<br>(0.021) | -0.0002<br>(0.021) | -0.004<br>(0.021) | -0.001<br>(0.021) | -0.003<br>(0.021) |
| Efficacy: 50% |  | 0.027<br>(0.024) | 0.027<br>(0.024) | 0.025<br>(0.024) | 0.023<br>(0.024) | 0.021<br>(0.024) | 0.020<br>(0.024) | 0.015<br>(0.024) |
| Insurance: medical |  | -0.047**<br>(0.020) | -0.048**<br>(0.020) | -0.049**<br>(0.020) | -0.048**<br>(0.020) | -0.042**<br>(0.020) | -0.043**<br>(0.020) | -0.042**<br>(0.020) |
| Insurance: medical + extra |  | -0.023<br>(0.020) | -0.023<br>(0.020) | -0.024<br>(0.020) | -0.022<br>(0.020) | -0.018<br>(0.020) | -0.017<br>(0.020) | -0.017<br>(0.020) |
| Mild: 0.1% |  | -0.034*<br>(0.021) | -0.036*<br>(0.021) | -0.036*<br>(0.021) | -0.038*<br>(0.021) | -0.035*<br>(0.021) | -0.035*<br>(0.021) | -0.034*<br>(0.020) |
| Mild: 1% |  | 0.009<br>(0.021) | 0.007<br>(0.021) | 0.007<br>(0.021) | 0.003<br>(0.021) | 0.004<br>(0.021) | 0.005<br>(0.021) | 0.008<br>(0.021) |
| Mild: 10% |  | -0.021<br>(0.022) | -0.023<br>(0.022) | -0.022<br>(0.022) | -0.024<br>(0.022) | -0.022<br>(0.022) | -0.019<br>(0.022) | -0.019<br>(0.022) |
| Severe: 0.0001% |  | -0.050**<br>(0.023) | -0.051**<br>(0.023) | -0.050**<br>(0.023) | -0.050**<br>(0.023) | -0.050**<br>(0.022) | -0.050**<br>(0.022) | -0.051**<br>(0.022) |
| Severe: 0.001% |  | -0.070***<br>(0.023) | -0.072***<br>(0.023) | -0.071***<br>(0.023) | -0.072***<br>(0.023) | -0.069***<br>(0.023) | -0.069***<br>(0.023) | -0.071***<br>(0.023) |
| Severe: 0.01% |  | -0.037<br>(0.024) | -0.038<br>(0.024) | -0.037<br>(0.024) | -0.042*<br>(0.024) | -0.036<br>(0.024) | -0.039*<br>(0.024) | -0.040*<br>(0.024) |
| Severe: No info |  | -0.022<br>(0.025) | -0.020<br>(0.025) | -0.018<br>(0.025) | -0.023<br>(0.025) | -0.023<br>(0.025) | -0.024<br>(0.025) | -0.024<br>(0.025) |
| Intention to be vaccinated x Tested |  |  |  |  |  |  |  | 0.151***<br>(0.032) |
| Tested x Confirmed case nearby (yes) |  |  |  |  |  |  |  | -0.133**<br>(0.056) |
| Tested x Confirmed case nearby (Not sure) |  |  |  |  |  |  |  | -0.0099<br>(0.075) |
| Constant | 1.262***<br>(0.046) | 1.344***<br>(0.054) | 1.322***<br>(0.056) | 1.323***<br>(0.069) | 1.210***<br>(0.073) | 1.243***<br>(0.079) | 1.244***<br>(0.079) | 1.344*** |
| Observations | 8,199 | 8,199 | 8,199 | 8,199 | 8,199 | 8,199 | 8,199 | 8,199 |
| Log Likelihood | -15,738 | -15,725 | -15,716 | -15,713 | -15,699 | -15,664 | -15,656 | -15,642 |
| Results (zero-inflated coefficients) from zero-inflated negative binomial regression |  |  |  |  |  |  |  |  |
| Intention to be vaccinated | -1.038***<br>(0.065) | -1.038***<br>(0.065) | -1.037***<br>(0.065) | -1.036***<br>(0.065) | -1.037***<br>(0.065) | -1.051***<br>(0.065) | -1.051***<br>(0.065) | -1.047***<br>(0.065) |
| Confidence in the vaccine | -0.310***<br>(0.020) | -0.310***<br>(0.020) | -0.311***<br>(0.020) | -0.311***<br>(0.020) | -0.311***<br>(0.020) | -0.301***<br>(0.020) | -0.300***<br>(0.020) | -0.301***<br>(0.020) |
| Constant | 1.944***<br>(0.079) | 1.944***<br>(0.079) | 1.944***<br>(0.079) | 1.943***<br>(0.079) | 1.944***<br>(0.079) | 1.929***<br>(0.079) | 1.929***<br>(0.079) | 1.926***<br>(0.079) |
| Observations | 8,199 | 8,199 | 8,199 | 8,199 | 8,199 | 8,199 | 8,199 | 8,199 |
| Log likelihood | -15,738 | -15,725 | -15,716 | -15,713 | -15,699 | -15,664 | -15,656 | -15,642 |

Note. \* p<0.1; \*\* p<0.05, \*\*\* p<0.01

Both of our predictors for the zero-inflation logit model – 1) intention to vaccinate and 2) confidence in the vaccine – are both statistically significant. Their negative estimates suggest that those who do not have the intention to be vaccinated and those who have lower confidence in the vaccine are more likely to be the “excess zeros” in the model, i.e. those who indicated 10 for time preference. Second, for the count model, our quickest observation is that vaccine attributes do not matter as much as respondent characteristics in determining vaccination time preference. Among the attributes, only severe side-effects (two of its levels, as compared to the smallest probability) and the provision of medical insurance (vs. no insurance provided) are statistically significant. However, the impact is not consistent across levels.

In comparison, respondent characteristics seem to matter more. To begin, all the demographic variables are statistically significant. Male, older, less educated, and higher income respondents prefer to be vaccinated earlier. Respondents who work in high risk occupations, including medical workers, service workers, janitors and frequent travellers, also prefer to be vaccinated earlier. Meanwhile, respondents who have higher stress levels, measured by the composite index of PHQ-4, are more likely to prefer being vaccinated earlier. However, there is no statistically significant relationship between having chronic illnesses, and residing with vulnerable persons (infants, chronic patients, pregnant women and elderly), and preferring to vaccinate earlier or later.

Expectedly, intention to vaccinate and confidence in the vaccines are both statistically significant predictors in the count model. Intention to be vaccinated, and higher confidence in the vaccines, contribute to earlier time preferences. Meanwhile, risk perceptions generally do not have statistically significant relationships with time preferences. Respondents who perceive the COVID-19 pandemic to be more serious, and those who think they have greater risk of being infected, are not more likely to want to be vaccinated earlier. However, respondents who think they are taking sufficient preventive measures are more likely to delay vaccination. Financial impact of the pandemic has no statistically significant relationship with vaccination time preference.

Our models also seek to test how various types of trust correlate with vaccination time preference. Similar to previous studies that trust in government increases vaccination intention (Lazarus *et al*., 2020), we also find that it has a statistically significant relationship with time preference. Respondents who have higher trust in government are more likely to prefer to be vaccinated earlier. Meanwhile, there is no statistical relationship between trust in medical workers and vaccination time preference. Important to note is that trust in friends has a negative statistical relationship with time preference. Respondents who trust their friends more tend to delay their vaccination.

Finally, whether a respondent has been COVID-tested is statistically related to vaccination time preference. Generally, respondents who have been recently tested are more likely to prefer later vaccination (Model 7). This makes sense because receiving a negative test result may give people a sense of protection, which lowers their guard against infection. At the same time, we also find that respondents who live in neighbourhoods where there are recently confirmed cases are more likely to prefer later vaccination. To understand this counterintuitive finding, we include two interaction terms – first between testing and intention to be vaccinated, and second between testing and the presence of confirmed cases nearby – into Model 8. This is to take into account the fact that people get tested for different reasons. Some may have gotten the test voluntarily; others may have been “forced” to take the test because of compulsion or lockdowns. We believe that different motivations for testing will shape time preference differently. The first interaction term is a proxy for whether testing is voluntary (if one wants to be vaccinated and has been tested, it is likely that the test is taken voluntarily). The second interaction term is a proxy for whether testing is involuntary (if one has been tested and there are confirmed cases nearby, it is likely that the test is involuntary).

The result shows that both interaction terms are statistically significant. Respondents who have been tested and have the intention to be vaccinated are more likely to prefer to be vaccinated *earlier* than those who have been tested with no such intention. In contrast, respondents who have been tested and are close to confirmed cases are more likely to be vaccinated later than those who have been tested but with no nearby confirmed cases. Meanwhile, after adding the second interaction term, the statistically significant effect for confirmed cases becomes insignificant, showing that whether there are confirmed cases around on its own will not affect time preference.

## Discussion

### Importance of efficacy and severe side-effect

With several COVID-19 vaccines being made available, public willingness to be vaccinated has become the next hurdle in fighting the pandemic. The results of our conjoint analysis showed that Hong Kong citizens are most concerned with vaccine efficacy and the likelihood of causing severe side effects when choosing COVID-19 vaccines. Although the importance of safety and efficacy have been identified previously in study in the US (Kreps *et al*., 2020; Motta, 2021), UK(McPhedran & Toombs, 2021) and China (Wang *et al*., 2020), our findings further contribute to these studies by showing that vaccines with 50% or 70% efficacy are 30.7% and 16.9% less likely to be chosen in comparison to a baseline vaccine with 95% efficacy, assuming all other attributes are comparable. This hypothetical situation means that if a country/region introduces vaccines with a great diversity of efficacy in the same mass vaccination program, citizens will be heavily inclined to choose the one with higher efficacy, potentially causing wastage of vaccines with a relatively lower efficacy.

We also found that the likelihood of causing severe side effects is a powerful contributor to preference for a vaccine. This result echoes previous research that indicates a more prominent role of adverse side-effect than mild side-effect in affecting public preference in other vaccines (de Bekker-Grob *et al*., 2010; Veldwijk *et al*., 2014; Guo *et al*., 2017). However, our results indicate that a vaccine with unknown probability of causing adverse side-effects is 8.1% less likely to be chosen than a vaccine with 0.01% chance of causing adverse effect. The finding suggests that information about the vaccines, especially their side-effects, should be made as transparent as possible, because uncertainty and low trust were prominent factors against timely vaccine uptake. Governments should enhance transparency of their vaccination program through effective communication.

### Value of medical insurance and free vaccine

In countries with well-established social welfare systems, COVID-19 vaccination programs may be accompanied by subsidy, indemnity and extra living support. For example, Japan and Hong Kong have enacted laws to make COVID-19 vaccines free to residents. The United Kingdom has set up vaccine damage payment to those injured by the COVID-19 vaccine. Singapore has also agreed to provide support to those having serious side effects after the jabs through applicable healthcare plans.

However, our study suggests that subsidy is a double-edged sword: people want their vaccine to be subsidized, but they will develop hesitancy if *too much* subsidy is offered. Results from the conjoint analysis indicate that subsidized vaccines are more preferable to a vaccine that costs money. However, respondents are more likely to choose a free vaccine over a subsidized vaccine. One explanation is that subsidies may generate a negative signaling effect that guides people to think about the negative consequences of receiving the vaccine. As the valuation of vaccine changes, people who place a higher value on reducing risk generated by vaccination will be less likely to receive vaccination (Cook *et al*., 2009). Furthermore, having reimbursement from vaccination is not common in other vaccination programs. The deviation from established practice may invoke unnecessary risk perception. This explanation may be applied to indemnity programs and other healthcare plans related to COVID-19 vaccination.

### Vaccination intention

Although up to 61% of respondents indicated a lack of intention to join the vaccine program, our findings show that their decision may change if they are presented with their preferred vaccine. Figure 2 suggests that while vaccination intention appears to correlate positively with time preference, there are more subtleties in the relationship. On the one hand, respondents who have the intention to be vaccinated may delay their time preference given a specific vaccine. On the other hand, although the majority of those who indicate no vaccination intention prefer to be vaccinated last regardless of the choice being presented, a significant proportion has chosen earlier time preferences. This implies that respondents who indicate no intention in being vaccinated may simply delay their timing. One possibility is that they are adopting a wait-and-see attitude to see whether the vaccines are safe enough. Another possibility is that rational people may want to avoid the potential cost of vaccination but benefit from herd immunity as public goods (Bauch *et al*., 2003; Bauch & Earn, 2004).

### Ensuring the success of vaccination programs

Effectiveness of a vaccination program can be jeopardized when a significant number of citizens are reluctant to receive the jabs early on. Vaccination programs should be planned meticulously to ensure a head start and avoid contention that may diminish public vaccination intention. For instance, identifying volunteers to join the early phase of vaccination programs can help ensure their smooth rollout and wider acceptance. Our time preference regression model suggests that demographic attributes are crucial predictors in vaccination time preference. Male, older, less educated, and higher income groups have higher tendency to be vaccinated earlier. Those who work in high-risk environments are also more likely to prefer receiving the jabs earlier than others. High stress level, as measured by PHQ-4 in this study, is another predictor to willingness to receive early vaccination. Vaccination programs should target citizens with these backgrounds as the priority in the early promotion of vaccination campaigns.

### Impact of trust

From the results, people who trust the government more tend to receive vaccination earlier, while people who trust their friends tend to delay their vaccination time and trust in medical workers have no relationship with vaccination time preference. Similar research has shown that individuals’ distrust or low trust against government is a significant predictor of vaccination hesitancy in other diseases (Quinn *et al*., 2013; Larson *et al*., 2018; Jamison *et al*., 2019). However, it remains largely unknown to us why individuals with higher trust in friends develop delay-vaccination sentiments. Extant literature shows that strong ‘bonding social capital’ (e.g., trust among friends) is associated positively with increasing vaccination intention and protective behaviour, which help create ‘public goods’ (e.g., herd immunity) under pandemic (Nagaoka *et al*., 2012; Rönnerstrand, 2013; Chuang *et al*., 2015). However, our results do not align with this finding. One possible explanation is that the distrust in government in Hong Kong undermines the benefit of state-led vaccination programs, leading to vaccination hesitancy and strong reliance on civil-society-led non-pharmaceutical interventions (Yuen *et al*., 2021). Another possibility is that people having better friendship networks may have some sorts of safety net to weather the uncertainties posed by the pandemic.

### Impact of forced COVID testing

Model 7 and 8 of our time preference analysis suggest that forced testing around the outbreak area may delay vaccination time preference, possibly owing to the fact that people may feel ‘safer’ after being tested and thus reduce the urgency to take the vaccine. Many governments are relying on forced testing and quarantine to contain the risk of an outbreak. However, previous studies have identified that the majority of COVID-19 infection is attributable to presymptomatic and asymptomatic infection, making forced testing inadequate to interrupt a COVID-19 outbreak unless contact tracing is made rapid and thorough (Moghadas *et al*., 2020). In addition, stringent public health policy may not entail policy compliance with COVID-19 related measures (Yue *et al*., 2021). Thus, the stringency, scale and timing of forced testing should be scaled up or down, depending on the severity of the outbreak and planning of the vaccine program.

## Conclusion

As more COVID-19 vaccines are becoming available, more countries are rolling out their vaccination plans. Understanding the factors that shape public preference allows public health officials to design better vaccination programmes and maximize vaccine uptake. In this study, citizens in Hong Kong express their preference for vaccines with both efficacy and safety. Our conjoint model also demonstrates the importance of medical insurance and free vaccines in promoting vaccination intention. The WHO’s guidance at this point lists out vaccine prioritization to high-risk populations. Our time preference analysis goes one step further by identifying the factors that determine when individuals prefer to be vaccinated. By highlighting the impact of trust and forced testing on vaccination time preference, we hope that our findings will inspire careful and creative vaccine planning.

Our study will have implications for different countries, whether or not they offer vaccine choices for citizens. For countries that offer only one type of vaccine, our findings suggest that the vaccination programme will proceed faster by offering a vaccine that promises a high enough efficacy and transparent information about the side-effects. After all, unless the cost of not-vaccinating is exceedingly high (e.g., being barred from bars and restaurants), people still have a ‘choice’ to delay their vaccination as much as possible. For countries where more than one vaccine option is available, our findings show that the public could be largely indifferent to vaccines with similar efficacy, odds of side-effects and insurance protection. Our experiment does not, however, exhaust all distinguishing attributes of COVID vaccines, and other studies have found, for instance, the country of origin of the vaccine (U.S. vs China vs Russia; Motta, 2021), as an additional attribute. Hence, we recommend public health scientists to be mindful of the possibility of other context-specific attributes that may influence vaccination choices.

Lastly, it is still uncertain how long the acquired immunity from COVID vaccines may last – i.e., whether COVID vaccine will require re-vaccination every now and then, or just a limited number of shots for life-long protection. There are likely to be more COVID vaccines entering the market in the foreseeable future, possibly in the form of a pill or nasal spray. COVID vaccination has received public engagement and attention unparalleled by most other vaccination programmes, and therefore offers public health a rare window to observe how the public learn, discuss and decide on receiving a brand-new vaccination. The learning we take from COVID vaccination could be generalized to reinvigorating other vaccination (e.g., HPV, influenza) and debunk anti-vaxxers’ myths.

## Data Availability

Data used in this research will be available upon request

## Funding Information

This research was funded by Innovative Research Grant awarded by the University of Hong Kong to [anonymized].

## Institutional Review Board Statement

The research has received ethics approval from the Human Research Ethics Committee of the University of Hong Kong under ref number EA2003003 (March 6, 2020).

## Informed Consent Statement

Informed consent was obtained from all subjects involved in the study.

## Conflict of Interest

The authors declare no conflict of interest.

## Appendix

### List of Table

**Table S1.** Summary descriptive statistics

| **Variable** | **Min** | **Max** | **Median** | **Mean** | **Standard deviation** |
| --- | --- | --- | --- | --- | --- |
| Age | 18 | 89 | 43 | 44.82 | 14.79 |
| Education | 1 | 6 | 4 | 4.17 | 1.40 |
| Sex | 0 | 1 | 1 | 0.32 | 0.47 |
| Income | 1 | 11 | 4 | 4.55 | 2.87 |
| Intention to vaccinate | 1 | 2 | 1 | 1.39 | 0.49 |
| Confidence in vaccine | 1 | 7 | 4 | 3.33 | 1.62 |
| High risk job | 0 | 1 | 0 | 0.34 | 0.47 |
| Chronic illness | 1 | 2 | 1 | 1.34 | 0.47 |
| Residing with vulnerable persons | 0 | 1 | 0 | 0.48 | 0.50 |
| Perception of the seriousness of COVID-19 | 1 | 10 | 8 | 7.25 | 1.89 |
| Perception of personal risk of being infected | 1 | 10 | 5 | 4.39 | 1.81 |
| Personal efforts to prevent being infected | 1 | 10 | 6 | 6.16 | 2.01 |
| Perceived financial impact caused by COVID-19 | 1 | 10 | 5 | 5.05 | 2.80 |
| Mental stress level (PHQ-4) | 4 | 16 | 6 | 7.02 | 2.95 |
| Trust in medical workers | 1 | 10 | 8 | 7.75 | 1.79 |
| Trust in the government | 1 | 10 | 3 | 3.50 | 2.68 |
| Whether the respondent has been tested | 0 | 1 | 1 | 0.52 | 0.50 |
| Whether there are confirmed cases near where the respondent lives in the past two weeks | 1 | 3 | 1 | 1.19 | 0.50 |

**Table S2:**
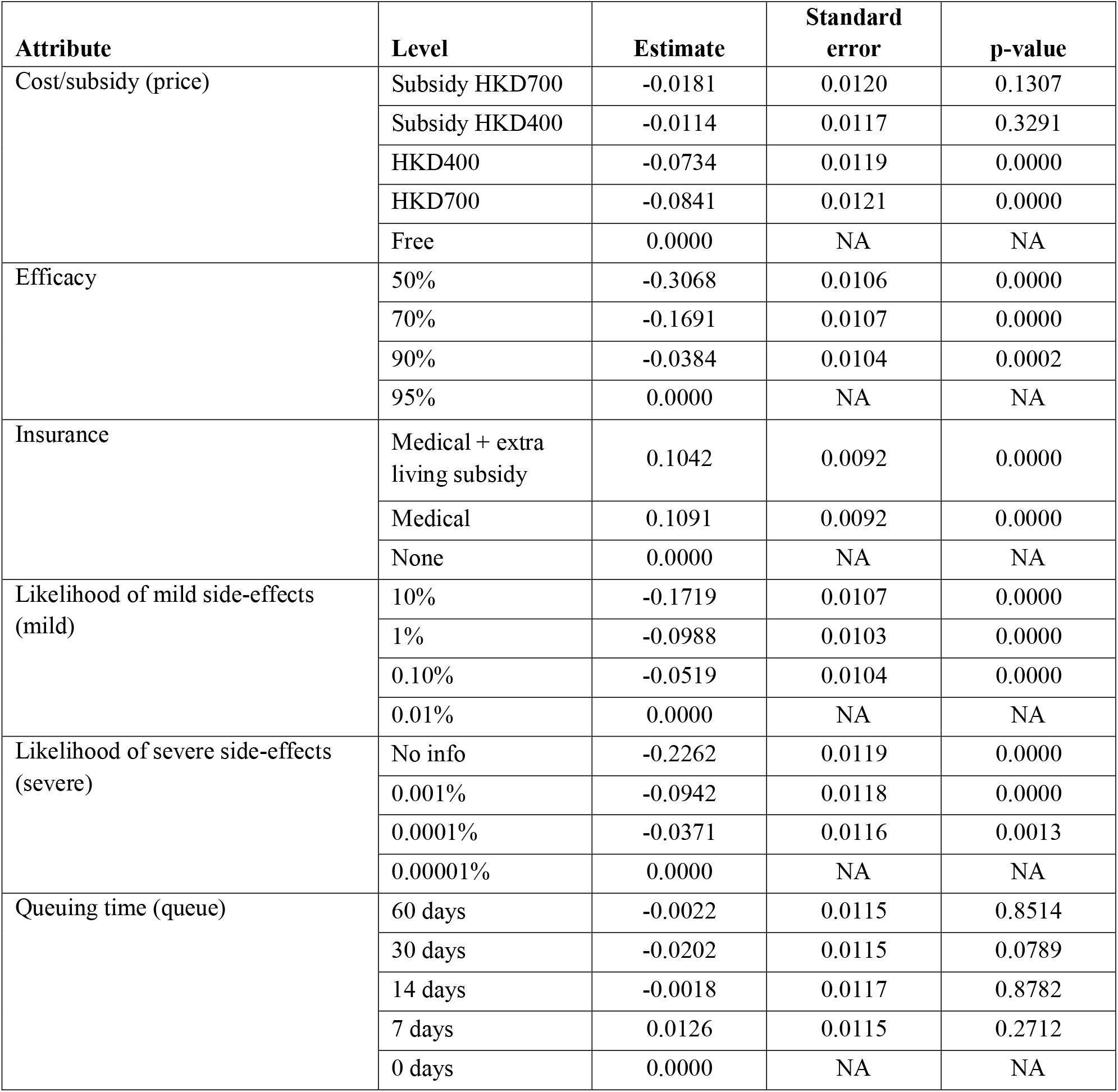
Coefficients of the conjoint model

| Attribute | Level | Estimate | Standard error | p-value |
| --- | --- | --- | --- | --- |
| Cost/subsidy (price) | Subsidy HKD700 | -0.0181 | 0.0120 | 0.1307 |
|  | Subsidy HKD400 | -0.0114 | 0.0117 | 0.3291 |
|  | HKD400 | -0.0734 | 0.0119 | 0.0000 |
|  | HKD700 | -0.0841 | 0.0121 | 0.0000 |
|  | Free | 0.0000 | NA | NA |
| Efficacy | 50% | -0.3068 | 0.0106 | 0.0000 |
|  | 70% | -0.1691 | 0.0107 | 0.0000 |
|  | 90% | -0.0384 | 0.0104 | 0.0002 |
|  | 95% | 0.0000 | NA | NA |
| Insurance | Medical + extra living subsidy | 0.1042 | 0.0092 | 0.0000 |
|  | Medical | 0.1091 | 0.0092 | 0.0000 |
|  | None | 0.0000 | NA | NA |
| Likelihood of mild side-effects (mild) | 10% | -0.1719 | 0.0107 | 0.0000 |
|  | 1% | -0.0988 | 0.0103 | 0.0000 |
|  | 0.10% | -0.0519 | 0.0104 | 0.0000 |
|  | 0.01% | 0.0000 | NA | NA |
| Likelihood of severe side-effects (severe) | No info | -0.2262 | 0.0119 | 0.0000 |
|  | 0.001% | -0.0942 | 0.0118 | 0.0000 |
|  | 0.0001% | -0.0371 | 0.0116 | 0.0013 |
|  | 0.00001% | 0.0000 | NA | NA |
| Queuing time (queue) | 60 days | -0.0022 | 0.0115 | 0.8514 |
|  | 30 days | -0.0202 | 0.0115 | 0.0789 |
|  | 14 days | -0.0018 | 0.0117 | 0.8782 |
|  | 7 days | 0.0126 | 0.0115 | 0.2712 |
|  | 0 days | 0.0000 | NA | NA |

## Footnotes

1 The Global Economic Outlook During the COVID-19 Pandemic: A Changed World, *The World Bank*, June 8 2020, https://www.worldbank.org/en/news/feature/2020/06/08/the-global-economic-outlook-during-the-covid-19-pandemic-a-changed-world.

## Notes

### Competing Interest Statement

The authors have declared no competing interest.

